# Characterizing Oseltamivir Use among Community-Dwelling Patients Diagnosed with Influenza Virus Infection, 2023-2025

**DOI:** 10.64898/2026.03.27.26349417

**Authors:** Emily A. McNair, Jennie H. Kwon, Carlos G. Grijalva, Son H. McLaren, Jessica E. Biddle, Samantha Dean, Lydia Bristol, Elizabeth B. White, Stephanie A. Fritz, Rachel M. Presti, Caroline A. O’Neil, Ellen Sano, Celibell Vargas, Jonathan Schmitz, Yuwei Zhu, Theresa A. Scott, Stacey House, H. Keipp Talbot, Melissa S. Stockwell, Alexandra M. Mellis, the RVTN Flu study group

## Abstract

**Background:** Oseltamivir is an antiviral medication for influenza that can reduce the duration of symptoms and may lower the risk of some complications. Recommendations for use of oseltamivir include in the outpatient setting for individuals at higher risk of developing influenza complications.

**Objectives:** To describe oseltamivir initiation and treatment completion among influenza-positive outpatients and identify factors associated with each.

**Methods:** In a U.S. outpatient household transmission study, index participants with laboratory-confirmed influenza provided up to 12 days of detailed information on medication use. We described oseltamivir initiation among index cases and treatment course completion of **≥** 10 doses among cases who initiated oseltamivir. We used unadjusted and adjusted logistic regression to identify factors associated with initiation and course completion.

**Results:** Among 823 enrolled index cases, 324 (39%) initiated oseltamivir treatment. Of 406 persons at higher risk for influenza complications, 172 (42%) initiated treatment. Oseltamivir initiation was lowest among children aged 2 to < 5 years (19%) compared to all other age groups. Among 313 cases who initiated oseltamivir, 42% completed the recommended treatment course of ≥10 doses. Among 163 individuals at higher risk of influenza complications, 69 (42%) completed the recommended treatment course of ≥ 10 doses. Children < 2 years were significantly less likely to complete treatment compared to adults aged 18-50 years (aOR: 0.21, 95% CI: 0.04, 0.78, p= 0.030); reasons for discontinuation could not be determined.

**Conclusions:** These findings reveal differences in oseltamivir treatment in an outpatient setting among groups at higher risk for influenza complications.

**Article Summary:** We described and analyzed oseltamivir initiation and course completion among influenza-positive outpatients enrolled in a household transmission study, finding low overall initiation and completion. Results highlight differences in antiviral treatment for groups at higher risk for influenza complications.

## 1. Introduction

Influenza contributes to a substantial burden of disease each year and can cause serious illness and complications among those infected [1]. Four antivirals are FDA-approved and currently recommended for treatment of influenza in the United States; oseltamivir is the most widely prescribed [2]. Oseltamivir is FDA approved for the treatment of influenza among individuals aged 14 days and older. The CDC [2] and American Academy of Pediatrics [3] recommend that oseltamivir should be started as soon as possible (< 48 hours of symptom onset) for people at higher risk of influenza complications [4] and recommends providers use clinical judgement for all others. Individuals at higher risk for influenza complications, or those who are hospitalized with influenza, may benefit from treatment initiation more than 48 hours after symptom onset [5]. The recommended course of oseltamivir for treatment of uncomplicated influenza is two doses per day for 5 days or 10 consecutive doses [2]. Among outpatients with mild influenza illnesses, oseltamivir may reduce symptom duration and hospital admission with most evidence suggesting a greater benefit when treatment is started within 48 hours after symptom onset [6-11].

Oseltamivir usage among outpatients varies by age and risk group. [12]. One investigation found that from 2010-2019, 37% of children under 2 years and 34% of children aged 2-5 years at higher risk of influenza complications and with an influenza diagnosis received prescriptions for oseltamivir [13]. A study of influenza-positive adults at higher risk for influenza complications treated in emergency department and urgent care settings enrolled in the VISION network during the 2023-2024 influenza season found that 58% were prescribed antivirals [14]. Once prescribed, taking an incomplete course of oseltamivir may reduce the impact treatment has on the course and severity of disease because current evidence is based on a 5 day or 10 dose course [2]. Currently there are few to no detailed data regarding course completion in the outpatient setting specific to oseltamivir in the United States. During the 2009 influenza H1N1 pandemic, global investigations reported rates of 79-88% for completing a five-day course (10 doses) of oseltamivir [5, 15-17]. However, findings during that period may not be representative of current US outpatient settings, where guidelines and dispensing patterns differ under non-pandemic influenza seasons.

The objective of our study is to provide a description and analysis of oseltamivir initiation and treatment completion in a population of influenza-positive individuals who presented for outpatient care and subsequently enrolled in a household transmission study.

## II. Methods

### Study design

As part of a case-ascertained household transmission investigation, we identified and enrolled individuals ranging in age from 0 to 96 years with laboratory-confirmed influenza infections. These individuals sought care at non-emergency ambulatory and emergency department settings in Tennessee, New York, and Missouri during the 2023-2024 and 2024-2025 influenza seasons. Index participants were defined as the first individuals within their households to test positive for influenza from a test performed by a health care provider. To be eligible for inclusion in the study, index participants had to have received a positive influenza test result within the previous five days, reside in a household with at least one other person, and have no household members who exhibited clinical symptoms suggestive of influenza on or preceding the date of symptom onset or the date of illness onset for the index case. Following provision of written consent, index participants were enrolled on study day 0, together with their household contacts, and prospectively followed for 7 days. For this analysis, we restricted our study population to index participants, since all received medical care and had the opportunity to be prescribed oseltamivir for treatment of influenza.

The study protocol was reviewed and approved as research by the Institutional Review Board at Washington University in St. Louis School of Medicine, with reliance from Columbia University Irving Medical Center, Vanderbilt University Medical Center, and CDC and was conducted consistent with applicable federal law and CDC policy.

### Data collection

Index participants self-reported sociodemographic information, including age, sex, and race and ethnicity at the time of enrollment. Participants also reported the presence of preexisting health conditions, including heart disease, asthma, cancer, diabetes, kidney, obesity, liver, and immune conditions. Self-reported influenza vaccination history was reviewed and verified by the study teams using available medical records, state immunization registries, and data from vaccine providers. Participants were considered vaccinated for the current season if they received a verified influenza vaccination or reported both a valid date and known location of vaccination. The date of vaccination had to occur at least 14 days prior to the onset date of the index case’s illness to be considered vaccinated. Participants were also instructed to self-collect nasal swab specimens daily during the 7-day follow-up period. Specimens were shipped to the central RVTN laboratory at Vanderbilt in Tennessee for processing and testing for influenza using the Hologic Panther Fusion™ quantitative RT-PCR instrument.

### Medication and Symptom Diaries

Index participants completed prospective and retrospective surveys designed to collect daily information on symptoms, medication usage, and medical care encounters. Retrospective daily surveys were conducted from the day prior to index symptom onset up until enrollment. After enrollment 7 days were assessed prospectively for a possibility of up to 12 days of data. Participants reported daily whether they had taken any over the counter and/or prescription medications including influenza antivirals (oseltamivir, baloxavir, peramivir, and/or zanamivir), and the number of doses taken for their current illness. In addition to medication use, participants reported daily any fever, chills, cough, sore throat, runny nose, nasal congestion, wheezing, shortness of breath, chest tightness, fatigue, muscle or body aches, loss of smell or taste, headache, abdominal pain, diarrhea, nausea, vomiting or any other symptom.

### Analysis

Index participants were included in analysis if they reported either “yes” or “no” to the question regarding the initiation of oseltamivir on at least 4 daily diaries. If we could not carry out last observation carried forward for participants missing initiation and dose data, they were excluded from the analysis. See the supplemental methods for more details. Among all included influenza-positive index cases, initiation of oseltamivir was defined as having ever taken 1 or more doses of oseltamivir throughout the course of the study. We described initiation across participant characteristics including characteristics that classify individuals as higher risk for influenza complications. In this analysis, higher-risk individuals were defined as people < 5 years (especially age < 2 years) or ≥ 65 years of age or people with heart disease, asthma, cancer, diabetes, kidney, obesity, liver, and immune conditions [4]. We also looked at the percentage of individuals at higher risk for influenza complications who initiated oseltamivir after seeking care more than 48 hours or 2 days after symptom onset because these individuals are recommended to initiate oseltamivir regardless of time to seek care. Finally, we performed unadjusted logistic regression using bidirectional Akaike information criterion (AIC) stepwise selection to select the final covariates for inclusion in our adjusted model for overall initiation. Variables controlled for in our adjusted models included season, sex, age, study enrollment site, care setting, preexisting conditions, and influenza vaccination.

To investigate oseltamivir course completion, we used a subset of the population who initiated oseltamivir. Participants were excluded from our population subset if their oseltamivir use was likely censored, defined as having reported taking oseltamivir on the first (one day prior to symptom onset or first baseline diary entry) or last day (last daily diary entry) of data collection. We defined a complete course of oseltamivir as 10 or more doses of oseltamivir to reflect clinical guidelines. We described each participant’s days of oseltamivir use, total doses of oseltamivir, dose distribution by age group, and whether their course was complete or incomplete. We did not ask participants to report reasons for oseltamivir discontinuation, however, among the subset of individuals who initiated oseltamivir, we conducted a supplemental analysis comparing symptoms at illness onset, treatment initiation, and treatment discontinuation between those who completed versus did not complete a full course of oseltamivir. Finally, we performed unadjusted and adjusted logistic regression for oseltamivir course completion among those who initiated treatment, controlling for season, sex, age, study enrollment site, care setting, preexisting conditions and influenza vaccination.

## III. Results

### Study Cohort

Of 890 index participants enrolled in the 2023-2024 and 2024-2025 seasons, 823 (92%) met inclusion criteria for the oseltamivir initiation analysis. (see Supplemental Figure 1). Of the total participants, 46% were enrolled during the 2023-2024 season and 54% were enrolled during the 2024-2025 season. Half (49%) of participants were at higher risk for influenza complications (Table 1). A majority of participants were children < 18, with 409/823 (50%) aged 5-17, 101/823 (13%) aged 2 to < 5 and 53/823 (6%) aged < 2. Of the 823 overall participants, 263 (32%) had preexisting medical conditions. Most (54%) were recruited from non-emergency ambulatory care settings and 36% had received a seasonal influenza vaccine (Table 1).

**Table 1.** Characteristics of influenza-positive index participants enrolled in the RVTN, stratified by analytic group **(n (%))**.

|  | Overall<br>N = 823 | Oseltamivir initiation<br>(N=823) |  | Oseltamivir completion<br>(N=313) |  |
| --- | --- | --- | --- | --- | --- |
|  |  | No<br>N = 499 | Yes<sup>1</sup><br>N = 324 | No (< 10<br>doses)<br>N = 180 | Yes (≥ 10<br>doses)<br>N = 133 |
| **Season** |  |  |  |  |  |
| 2023-2024 | 376 (46%) | 256 (68%) | 120 (32%) | 56 (50%) | 56 (50%) |
| 2024-2025 | 447 (54%) | 243 (54%) | 204 (46%) | 124 (62%) | 77 (38%) |
| **Sex<sup>2</sup>** |  |  |  |  |  |
| Female | 444 (54%) | 278 (63%) | 166 (37%) | 96 (59%) | 67 (41%) |
| Male | 372 (45%) | 217 (58%) | 155 (42%) | 83 (56%) | 64 (44%) |
| **Age (years)** |  |  |  |  |  |
| < 2 | 53 (6%) | 31 (58%) | 22 (42%) | 19 (86%) | 3 (14%) |
| 2-4 | 101 (13%) | 82 (81%) | 19 (19%) | 8 (44%) | 10 (56%) |
| 5-17 | 409 (50%) | 272 (67%) | 137 (33%) | 77 (60%) | 51 (40%) |
| 18-50 | 176 (21%) | 89 (51%) | 87 (49%) | 42 (49%) | 44 (51%) |
| 51-64 | 60 (7%) | 17 (28%) | 43 (72%) | 23 (53%) | 20 (47%) |
| ≥ 65 | 24 (3%) | 8 (33%) | 16 (67%) | 11 (69%) | 5 (31%) |
| **Race-Ethnicity** |  |  |  |  |  |
| American Indian and<br>Alaska Native, Non-Hispanic | 4 (1%) | 2 (50%) | 2 (50%) | 1 (50%) | 1 (50%) |
| Asian, Non-Hispanic | 11 (1%) | 4 (36%) | 7 (64%) | 5 (71%) | 2 (29%) |
| Black, Non-Hispanic | 125 (15%) | 82 (66%) | 43 (34%) | 23 (56%) | 18 (44%) |
| Hispanic/Latino | 280 (34%) | 198 (71%) | 82 (29%) | 46 (61%) | 30 (39%) |
| Multiple race, Non-Hispanic | 8 (1%) | 2 (25%) | 6 (75%) | 4 (67%) | 2 (33%) |
| White, Non-Hispanic | 355 (43%) | 183 (52%) | 172 (48%) | 94 (56%) | 75 (44%) |
| Unknown/Refused | 13 (2%) | 7 (54%) | 6 (46%) | 4 (67%) | 2 (33%) |
| Missing | 27 (3%) | 21 (78%) | 6 (22%) | 3 (50%) | 3 (50%) |
| **Site** |  |  |  |  |  |
| New York | 289 (35%) | 216 (75%) | 73 (25%) | 44 (66%) | 23 (34%) |
| Tennessee | 243 (30%) | 105 (43%) | 138 (57%) | 75 (56%) | 60 (44%) |
| Missouri | 291 (35%) | 178 (61%) | 113 (39%) | 61 (55%) | 50 (45%) |
| **Index Screening Site** |  |  |  |  |  |
| Non-emergency<br>ambulatory care<sup>3</sup> | 443 (54%) | 220 (50%) | 223 (50%) | 123 (56%) | 98 (44%) |
| Emergency care | 380 (46%) | 279 (73%) | 101 (27%) | 57 (62%) | 35 (38%) |
| **Preexisting medical**<br>**conditions** |  |  |  |  |  |
| No underlying condition<sup>4</sup> | 552 (67%) | 364 (66%) | 188 (34%) | 109 (60%) | 73 (40%) |
| Underlying condition<sup>5</sup> | 263 (32%) | 130 (49%) | 133 (51%) | 71 (55%) | 57 (45%) |
| Missing/Unknown | 8 (1%) | 5 (63%) | 3 (37%) | 0 (0%) | 3 (100%) |
| **At higher risk for influenza complications** |  |  |  |  |  |
| No | 411 (50%) | 261 (64%) | 150 (36%) | 83 (57%) | 62 (43%) |
| Yes<sup>6</sup> | 406 (49%) | 234 (58%) | 172 (42%) | 97 (58%) | 69 (42%) |
| Missing/Unknown | 6 (1%) | 4 (67%) | 2 (33%) | 0 (0%) | 2 (100%) |
| **Influenza vaccination status** |  |  |  |  |  |
| Unvaccinated | 512 (62%) | 335 (65%) | 177 (35%) | 100 (60%) | 68 (40%) |
| Vaccinated<sup>7</sup> | 286 (35%) | 149 (52%) | 137 (48%) | 75 (55%) | 61 (45%) |
| Missing/Unknown | 25 (3%) | 15 (60%) | 10 (40%) | 5 (56%) | 4 (44%) |
| **Symptom onset to care seeking date (days)** |  |  |  |  |  |
| 0 days | 82 (10%) | 42 (51%) | 40 (49%) | 21 (55%) | 17 (45%) |
| 1-2 days | 503 (61%) | 280 (56%) | 223 (44%) | 126 (58%) | 92 (42%) |
| 3-4 days | 220 (27%) | 163 (74%) | 57 (26%) | 31 (57%) | 23 (43%) |
| 5-6 days | 18 (2%) | 14 (78%) | 4 (22%) | 2 (67%) | 1 (33%) |
| **Symptom onset to oseltamivir initiation (days)** |  |  |  |  |  |
| 0 days | 29 (4%) | 0 (0%) | 29 (100%) | 11 (41%) | 16 (59%) |
| 1-2 days | 188 (23%) | 0 (0%) | 188 (100%) | 101 (55%) | 84 (45%) |
| 3-4 days | 77 (9%) | 0 (0%) | 77 (100%) | 47 (62%) | 29 (38%) |
| 5-6 days | 21 (3%) | 0 (0%) | 21 (100%) | 15 (79%) | 4 (21%) |
| 7+ days | 9 (1%) | 0 (0%) | 9 (100%) | 6 (100%) | 0 (0%) |

<sup>1</sup>Initiation is defined as any participant who took $\geq 1$ dose of oseltamivir throughout follow-up.
<sup>2</sup>Overall, 7 out of 823 participants (1%) did not report their sex. Of these, 4 (57%) did not initiate oseltamivir, while 3 (43%) did. Among those who initiated oseltamivir, 1 (33%) took < 10 doses and 2 (67%) took $\geq 10$ doses of oseltamivir.
<sup>3</sup>Ambulatory care settings include walk-in clinics, pharmacies, occupational health, and specialized care clinics.
<sup>4-5</sup>Underlying conditions include heart disease, asthma, cancer, diabetes, kidney, obesity, liver, and immune conditions.
<sup>6</sup>At higher risk for influenza complications is defined as anyone < 5 years of age (especially age < 2 years), $\geq 65$ years of age, or who have a preexisting condition defined as increasing the risk for influenza complications [4].
<sup>7</sup>Participants were considered vaccinated for influenza if they received a verified influenza vaccination or if a valid date and location of vaccination were reported at least 14 days prior to the onset date of the index case's illness.

### Oseltamivir Initiation

Overall, 324 out of 823 participants (39%) reported initiating oseltamivir (Table 1). Among other influenza antivirals, baloxavir was initiated by 4 participants (one of whom also reported oseltamivir use) and zanamivir by 1 participant. These participants were retained in the analysis. Among all participants who are at higher risk for influenza complications, 172/406 (42%) initiated oseltamivir (Table 1). Oseltamivir initiation was higher among children aged < 2 (42%, 22 of 53) compared to children aged 2 to < 5 (19%, 19 of 101). In contrast, adults aged ≥ 65 had a higher initiation rate of 67% (16 of 24, Table 1). Among 263 participants with preexisting conditions (including any of: heart disease, cancer, diabetes, obesity, and various liver, kidney, lung, and immune conditions), 133 (51%) initiated oseltamivir compared to 34% with no preexisting conditions (Table 1). Pre-existing conditions were more common among older adults than children, and frequencies of initiation stratified by both age and underlying conditions are given in Supplemental Table 1. Across study sites, initiation was highest in Tennessee (57%), and more (50%) participants screened and enrolled from non-emergency ambulatory care settings initiated oseltamivir compared to those from emergency care settings (27%) (Table 1).

Across all index participants, the median duration from symptom onset to seeking care was 2 days (interquartile range (IQR): 0–3 days) and a majority of participants initiated oseltamivir on the day they sought care, as detailed in Supplemental Table 2. Initiation of oseltamivir was more common among the 585 overall participants who sought care within the first 48 hours after symptom onset (45%) compared to the 237 overall participants who sought care more than 48 hours after symptom onset (26%) (see Supplemental Table 2 and Table 3). This was also true among individuals at higher risk for influenza complications recommended to receive oseltamivir, with a consistent pattern across all age groups and presence of preexisting medical conditions (see Supplemental Table 3).

### Odds of Oseltamivir Initiation

In the unadjusted analysis, participants at higher risk for influenza complications based on age and preexisting conditions were 1.28 times more likely to initiate oseltamivir, but the association was not significant (p > .05) (Figure 1). Ages 2 to < 5 were associated with decreased odds of oseltamivir initiation compared to adults aged 18-50 (OR: 0.24, 95% CI: 0.13, 0.42, p < 0.001), while the presence of preexisting conditions was associated with increased odds of oseltamivir initiation (OR: 1.98, 95% CI: 1.47–2.67, p < 0.001) compared to those without preexisting conditions (Figure 1) Adjusted analyses did not include overall higher-risk status but instead included age and underlying medical conditions as individual covariates. In these adjusted analyses children aged 2 to < 5 were significantly less likely to initiate oseltamivir than adults aged 18-50 (aOR: 0.30, 95% CI: 0.15, 0.58, p < 0.001) (Figure 1). Additionally, participants enrolled from Tennessee exhibited higher odds of initiating oseltamivir compared to those from New York (aOR: 2.41, 95% CI: 1.32, 4.38, p = 0.008) (Figure 1).

**Figure 1.**
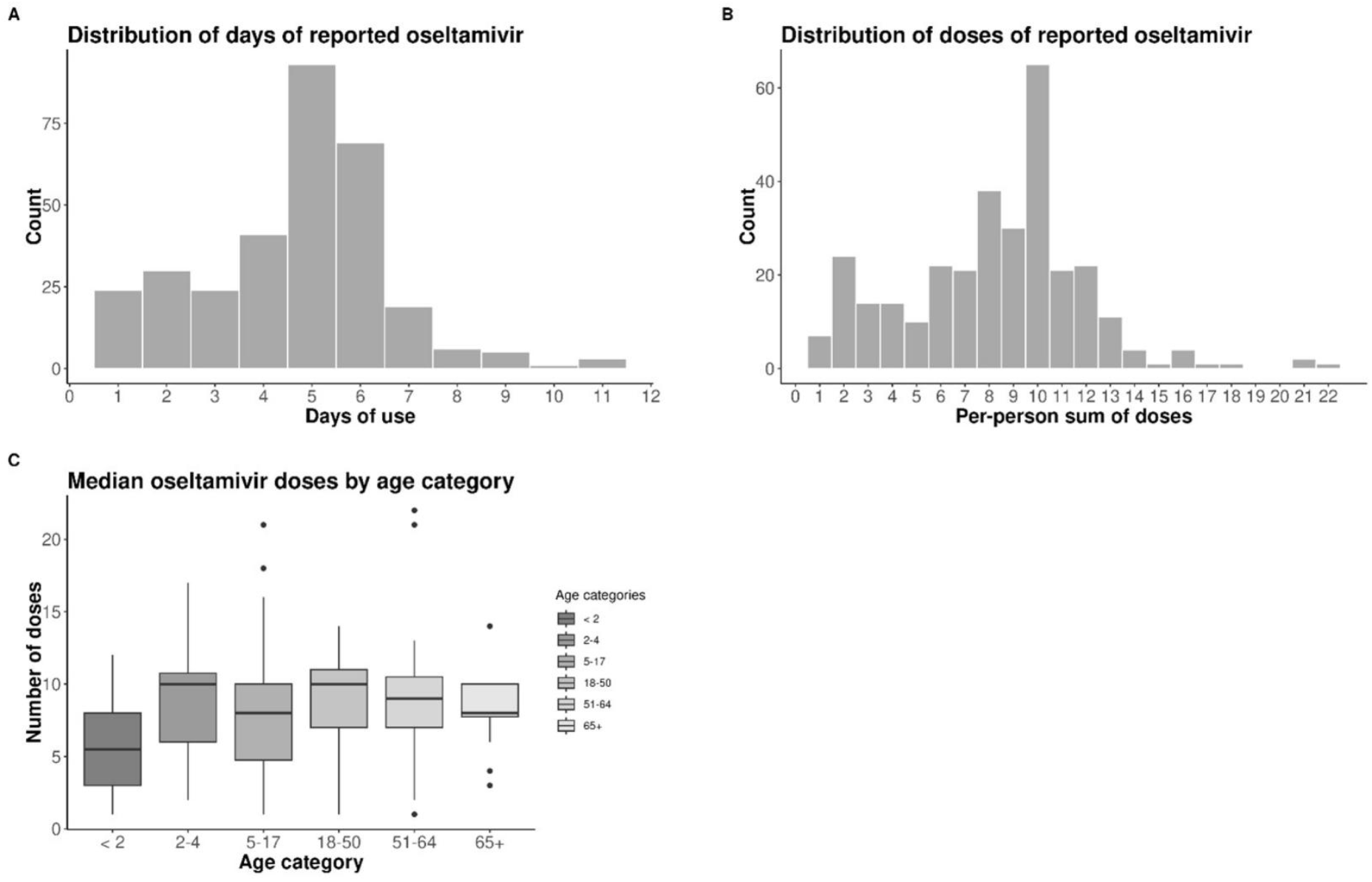
Descriptions of oseltamivir initiation and dosages in the outpatient cohort of influenza-positive index participants enrolled in the Respiratory Viral Transmission Network. **Panels A, B, and C:** Histograms and box plot illustrating the use of oseltamivir among participants who initiated treatment, excluding those with likely censored usage. **Panel A:** Histogram displaying the distribution of days of oseltamivir use, with a maximum of 12 days of data collection available. **Panel B:** Histogram depicting the distribution of oseltamivir doses. **Panel C:** Box plots representing the median doses of oseltamivir stratified by age group.

### Oseltamivir Course Completion

The number of days of oseltamivir use ranged from 1 day to 11 days and the median number of days of use was 5 for all enrolled index participants (IQR: 4-6 days) (Figure 1, Panel A). The number of oseltamivir doses taken by all participants ranged from 1 dose to 22 doses and the median number of doses taken was 9 (IQR: 6-10 doses) (Figure 1, Panel B). The median number of oseltamivir doses taken was lowest among participants < 2 years of age (median: 5, IQR: 3-8 doses) (Figure 1, Panel C).

We described oseltamivir course completion among a subset of 313 index participants whose oseltamivir use was not censored by the beginning or end of follow-up (see Supplemental Figure 1). To align with clinical guidelines, we defined completion as receiving ≥ 10 doses of oseltamivir. Overall, 42% of participants completed the recommended course (Table 1). Participants at risk for influenza complications completed a course of ≥ 10 doses at the same rate as the overall population (42%) (Table 1). Completion of a ≥ 10 dose course was lowest among children aged < 2 years (25%) and adults aged 65 years and older (31%) while higher among children aged 2 to < 5 years (56%) (Table 1). Additionally, 45% of individuals with preexisting conditions completed a ≥ 10 dose course (Table 1). A greater proportion of participants who initiated oseltamivir on the day of symptom onset completed a ≥ 10 dose course compared to the proportion of participants who initiated oseltamivir 1-7+ days after symptom onset (Table 1). The proportion of participants reporting symptoms was similar at illness onset, treatment initiation, or treatment discontinuation between individuals who completed versus did not complete a ≥ 10 dose course. (see Supplemental Figure 3).

### Odds of Oseltamivir Completion

In unadjusted analysis, participants at higher risk for influenza complications and recommended to receive oseltamivir had 0.95 odds of completing a ≥ 10 dose course compared to those not at higher for influenza complications, but the results were not significant (p > .05). Individuals aged < 2 years had significantly lower odds of completing a ≥ 10 dose course of oseltamivir in the unadjusted model compared to those aged 18-50 years (OR: 0.15, 95% CI: 0.03, 0.48, p= 0.004) and in the adjusted model (aOR: 0.21, 95% CI: 0.04, 0.78, p= 0.030). No additional variables including the prespecified variables for selection in the model, season, sex, age, presence of preexisting conditions, study site, care setting, or influenza vaccination status reached statistical significance in either the unadjusted or adjusted analyses. (Figure 2).

**Figure 2.**
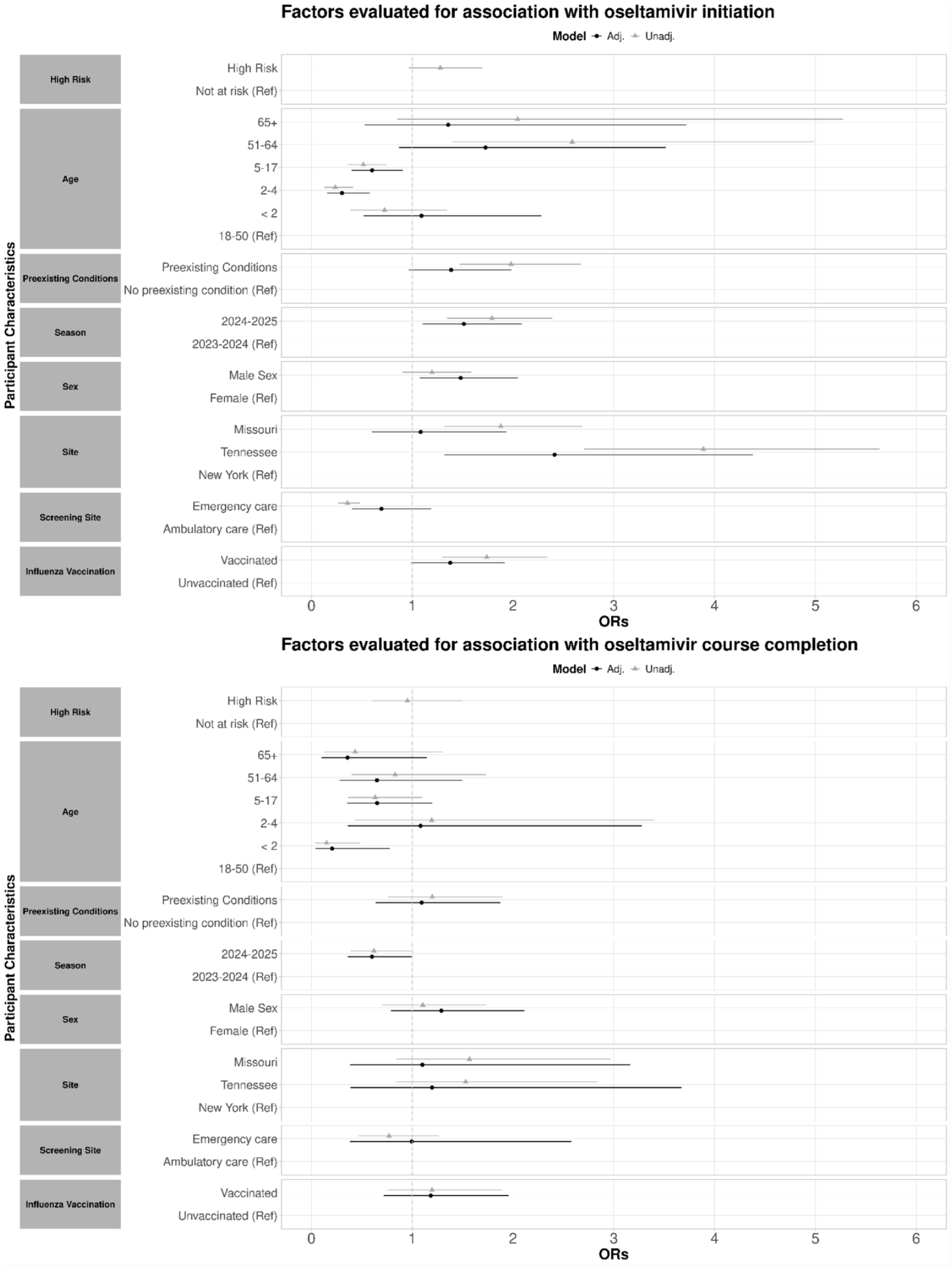
Unadjusted and adjusted odds ratios of oseltamivir initiation and completion by select characteristics in our outpatient cohort of influenza-positive index participants enrolled in the Respiratory Viral Transmission Network. Results of logistic regression for oseltamivir treatment outcomes: initiation (defined as taking at least one dose) and course completion (defined as completing a full course of at least 10 doses). All estimates except for higher-risk status were adjusted for season, sex, age, recruitment site, screening site (care setting), preexisting conditions, and vaccination status. Estimates with confidence intervals overlapping 1 are considered non-significant.

## IV. Discussion

This report describes oseltamivir initiation and course completion among people of all ages with laboratory-confirmed influenza infection enrolled in an outpatient household transmission study. Identifying patterns in oseltamivir initiation and course completion enhances our understanding of outpatient antiviral use in the United States. A minority of overall participants (39%) initiated oseltamivir. Among those classified as higher risk for influenza complications, less than half (42%) initiated oseltamivir, compared to 36% of participants not considered higher risk. However, we found that some higher-risk groups are less likely to initiate oseltamivir than others. Only 19% of participants aged 2 to < 5 years initiated oseltamivir while initiation was greater among participants aged **≥** 65 years (67%). We additionally found that overall, fewer than half of people who initiated oseltamivir completed a **≥** 10 dose course, representing the recommended twice daily dosing for 5 days. Individual courses varied, but the lowest median number of doses taken was among children at higher risk for influenza complications aged < 2 years.

The 2024-2025 influenza season was classified as high severity for all ages, with high hospitalization rates among children [18], a high number of pediatric deaths [19], and reported cases of influenza associated encephalopathy (IAE), a rare but serious neurologic influenza complication, in children [20]. Despite severe influenza seasons, outpatient investigations have found low initiation of oseltamivir, particularly among children, beyond the present report. One investigation using outpatient and emergency department prescription claims included in the IBM Marketscan Commercial Claims and Encounters Database, found that from 2010-2019, 37% of children < 2 of age and 34% of children 2-5 years of age with an influenza diagnosis received antiviral treatment [13].

Although we did not collect data on prescribing practices, several factors can influence whether medical professionals prescribe antivirals to groups at higher risk for influenza complications, including children. These include providers’ perceptions of the risks and benefits for each patient, as well as concerns about potential side effects, which have been reported in provider surveys [21]. Additional factors may influence whether participants choose to initiate oseltamivir, such as concerns about potential side effects or a perceived lack of illness or risk for severe disease [22]. While we did not capture these factors in our analysis, others such as time to seek care were found to be related to antiviral initiation in our analysis. Data from the Influenza Vaccine Effectiveness Network also found that early presentation to care was significantly associated with receipt of an influenza antiviral prescription [23], likely reflecting provider adherence to guidelines recommending treatment initiation within 48 hours of symptom onset. We also captured differences in initiation across settings, finding higher initiation among participants enrolled from Tennessee. Geographic differences may reflect oseltamivir availability or differences in provider knowledge. Our findings highlight the heterogeneity of oseltamivir initiation in the outpatient setting, highlighting the difference between current recommendations and real-world use.

This study is among the first to describe individual courses of oseltamivir in the outpatient setting, and found that among the individuals that initiated oseltamivir, not all completed a recommended course of ≥ 10 doses. Fewer than half of participants with preexisting conditions completed a full course and children < 2 years of age had the lowest rate of course completion. Although evidence from a systematic review found oseltamivir to be associated with gastrointestinal side effects, especially for pediatric age groups [24], we did not observe significant differences in reported gastrointestinal symptoms at illness onset, treatment initiation, or treatment discontinuation between individuals who completed and those who did not complete oseltamivir courses. These data could not confirm that gastrointestinal side effects drive early discontinuation. We also did not observe differences in other reported symptoms and could not confirm that symptom resolution drove discontinuation. While we did not collect any additional information from participants relating to reasons for discontinuation, other factors such as perceptions of antiviral effectiveness and counseling by healthcare providers, specifically pharmacists, may substantially impact treatment completion [25, 26]. As is currently recommended, timely initiation of a 5-day treatment course or **≥** 10 doses of oseltamivir maximizes the benefit of oseltamivir but was not completed by a majority of our study participants.

Our study is subject to several limitations. First, data on oseltamivir use was self-reported, which may introduce bias into our study. Second, participants enrolled in the study were not asked to report whether they received a prescription for an antiviral, so the number of people receiving a prescription may have been different from the number who ultimately initiated oseltamivir. Third, precision of our estimates was limited due to small cell sizes, particularly within pediatric age groups. Additionally, our findings are likely not representative as we only enrolled participants who sought care from three U.S academic medical centers and had been given an influenza test and ultimately agreed to participate in a prospective study with intensive daily follow-up.

This study describes the initiation and completion of a course of 10 or more doses of oseltamivir among influenza-positive participants presenting for outpatient care across three US sites. We found low overall rates of both initiation and completion of a ≥ 10-dose course of oseltamivir. Initiation was lowest among pediatric participants, specifically ages 2 to < 5 years, while course completion was lowest for ages < 2 years. Improved understanding of oseltamivir initiation and treatment completion for influenza in populations at higher risk for influenza complications can aid in the evaluation of clinical outcomes, which in turn helps determine whether there is a need for improving prescribing patterns for outpatients. However, given the high burden of influenza among patients at higher risk for influenza complications, antiviral treatment and course completion remain important.

## Data Availability

Data are available upon reasonable request.

## Acknowledgements

Thank you to the RVTNFlu Study Group Members: Kim T. Vu, Regine Burton, Francesca Yerbic, Lucy Vogt, Carleigh Samuels, Kelly Bono, Olivia Arter, Alyssa Valencia, Elianora Ovchiyan, Akshay Saluja, Mengesha Teshome, Alexandra Astarita, Sara Hubert, Tina Robinson, Mary G. Boyle, Alaina L. Schneider, Lisa M. Richardson, Tara Curley, Jamie Mills, Elizabeth Capasso-Gulve, Sarah Pruitt-Welsh, Sam Kerz, Heena Kahn, Jamie Garner, Judy A. King, Erica Anderson, Brianna Schibley-Laird, Rebecca Tablada, Onika Abrams, Sydney A. Cornelison, Dandan Liu, Liqun Wang, Anny L Diaz Perez, Raul A. Silverio Francisco, Ana Valdez de Romero

