## Supplemental Materials for "Characterizing Oseltamivir Use among Community-Dwelling Patients Diagnosed with Influenza Virus Infection, 2023-2025"

**Supplemental methods**

**Imputation of missing responses for daily oseltamivir use**: Only participants with some incomplete medication information were included in this analysis. Individuals answered, “Have you/has your child taken any medication to treat your/their symptoms?” during the 2023-2024 influenza season, and “On [calendar date] did [participant] take any prescription medicine for cold or flu symptoms, fever, and/or pain?” during the 2024-2025 influenza season. Participants had the option to respond “yes”, “no”, or “unsure” for oseltamivir initiation on each day of the daily and baseline diaries. If they answered “yes” participants were then asked to report doses of oseltamivir taken that day. Answer options included “1 dose”, “2 doses” or “unsure”. For participants who responded "unsure" about oseltamivir initiation on a single day, we applied last observation carried forward (LOCF) imputation for oseltamivir initiation, assuming medication use remained unchanged since the last recorded entry. If we could not carry out LOCF for participants, they were excluded from the analysis. If one or more response was blank (no answer provided) they were considered as having “no” oseltamivir initiation for that day. There were no blank responses adjacent to any reported initiation days. We additionally applied LOCF imputation to reported oseltamivir doses.

**Supplemental results**

**Supplemental Figure 1.** Creation of study groups.

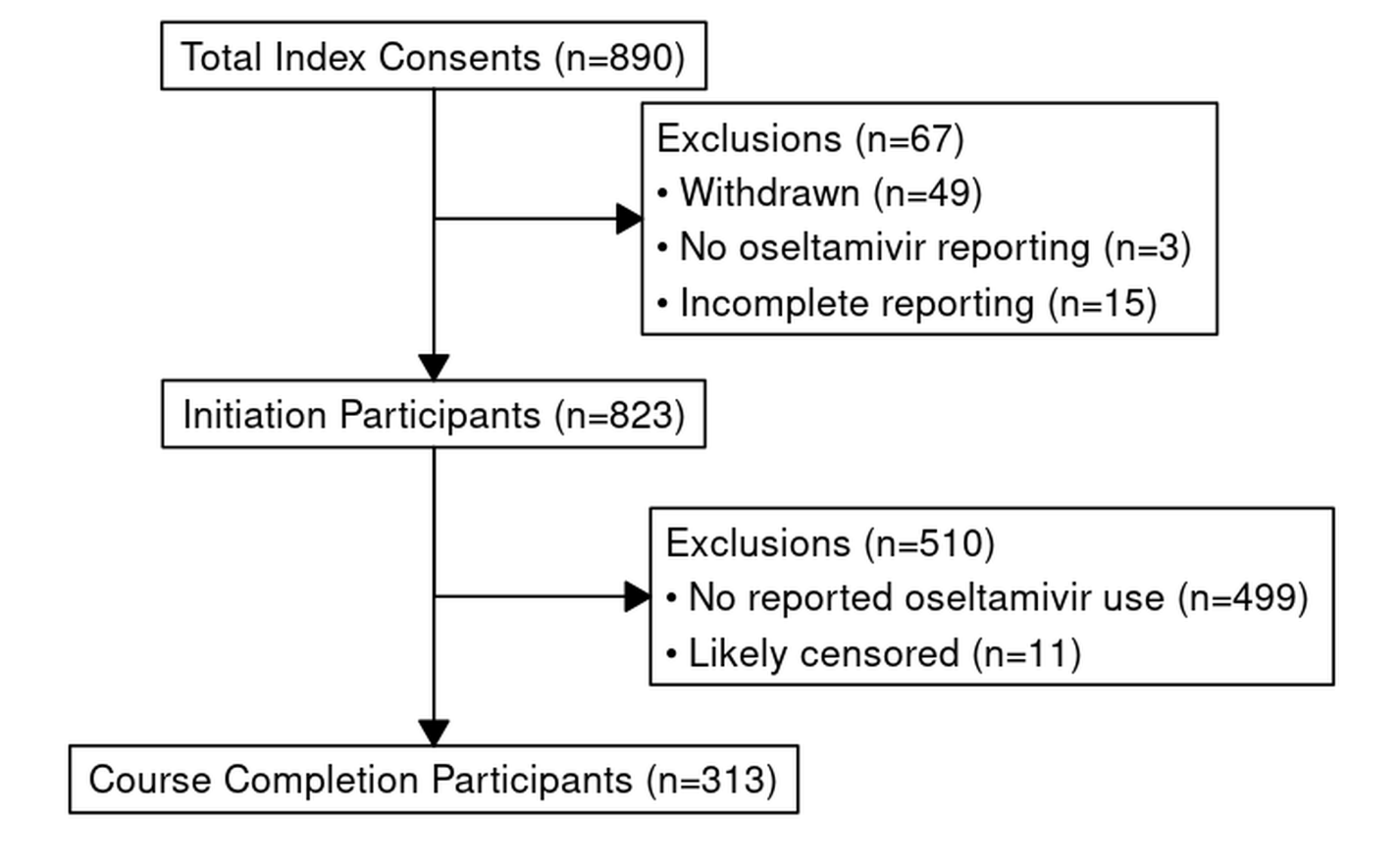

**Supplemental Figure 2.** Distributions describing time from symptom onset to care seeking date, time from care seeking date to oseltamivir initiation, and time from symptom onset to oseltamivir initiation in our outpatient cohort of influenza-positive index participants enrolled in the Respiratory Viral Transmission Network.

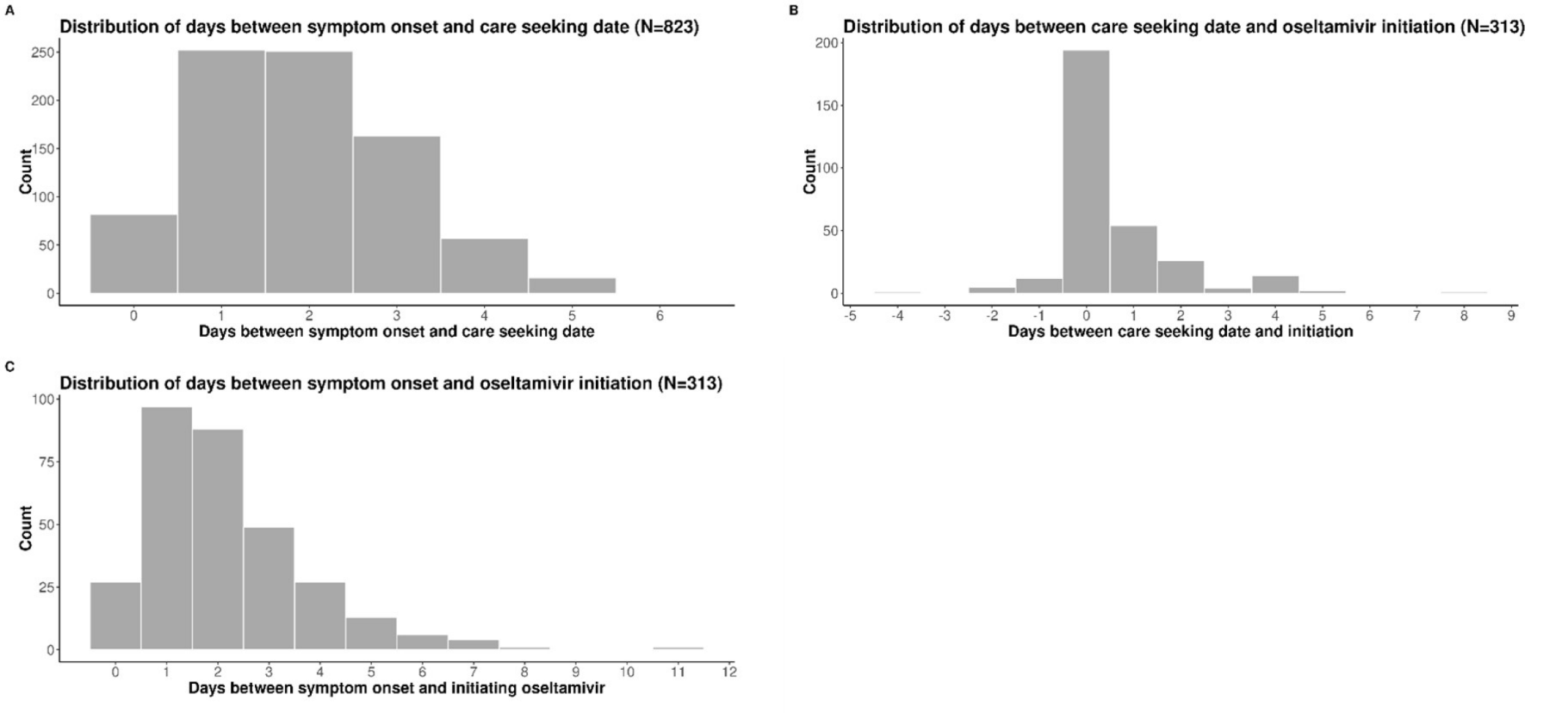

**Panel A:** Histogram of days between symptom onset and care seeking date, out of index participants who did not withdraw from the study and had complete oseltamivir reporting. **Panel B:** Histogram of days between care seeking date and date of first oseltamivir dose, out of those who initiated treatment, excluding those with likely censored usage. **Panel C:** Histogram of days between symptom onset and initiating oseltamivir, out of those who initiated treatment, excluding those with likely censored usage.

Sy===

**Supplemental Figure 3.** Proportion of study participants reporting symptoms on the day of influenza onset and on the day of oseltamivir initiation, stratified by course completion (≥ 10 vs < 10 doses) in our outpatient cohort of influenza-positive index participants enrolled in the Respiratory Viral Transmission Network.

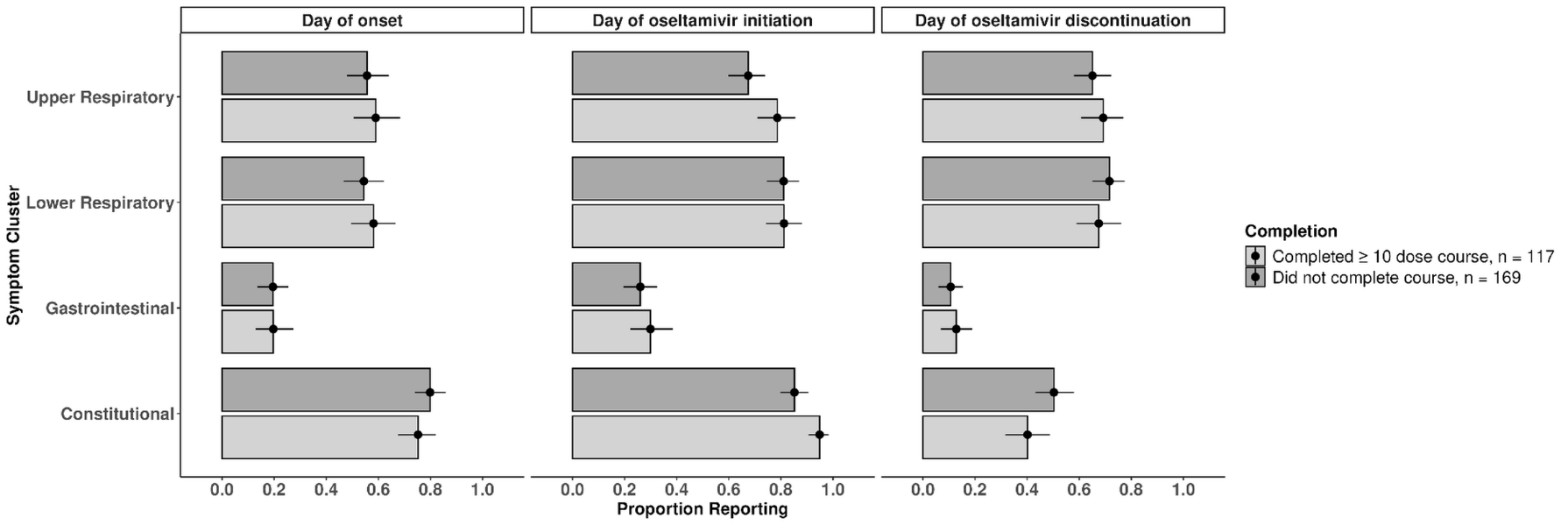

Symptoms were classified as: Upper respiratory (sore throat, runny nose, or nasal congestion); Lower respiratory (cough, wheezing, shortness of breath, or chest tightness/pain); Gastrointestinal (abdominal pain, diarrhea, or vomiting) or Constitutional (fever, aches, or fatigue).  This figure excluded 38 persons whose oseltamivir use was initiated on the same day as onset. Data were analyzed using logistic regression for comparison of symptoms between individuals who completed treatment and those who did not, with no significant associations between treatment completion and symptoms reported (regression results not shown).

**Supplemental Table 1.** Participants stratified by presence of preexisting conditions reporting on whether they initiated oseltamivir, by age category in our outpatient cohort of influenza-positive index participants enrolled in the Respiratory Viral Transmission Network.

|  | No preexisting condition N= 552 | | | Preexisting condition^1^ N= 263 | | |
| --- | --- | --- | --- | --- | --- | --- |
| Age category^2^ | **Overall** | **No oseltamivir (%)** | **Initiated oseltamivir (%)** | **Overall** | **No oseltamivir (%)** | **Initiated oseltamivir (%)** |
| < 5 | 137 (25%) | 102(75%) | 35 (25%) | 15 (5%) | 10 (68%) | 5 (32%) |
| 5-64 | 411 (74%) | 261 (64%) | 150 (36%) | 228 (87%) | 113 (49%) | 115 (51%) |
| ≥ 65 | 4 (1%) | 1 (25%) | 3 (75%) | 20 (8%) | 7 (35%) | 13(65%) |

^1^Underlying conditions include heart disease, asthma, cancer, diabetes, kidney, obesity, liver, and immune conditions.

^2^Age categories included reflect the stratification of children and adults, where children <5 (especially age < 2 years) are at higher risk of influenza complications [4].

**Supplemental Table 2.** Time to seek care by overall risk status, age category, and preexisting conditions in our outpatient cohort of influenza-positive index participants enrolled in the Respiratory Viral Transmission Network.

|  | Sought care within 48 hours of symptom onset | Did not seek care within 48 hours of symptom onset |
| --- | --- | --- |
| Overall | 585 (71%) | 237 (29%) |
| At higher risk for influenza complications |  |  |
| No | 304 (74%) | 107 (26%) |
| Yes^1^ | 278 (69%) | 127 (31%) |
| Missing/Unknown | 3 (50%) | 3 (50%) |
| Age categories |  |  |
| < 2 | 42 (81%) | 10 (19%) |
| 2-4 | 65 (64%) | 36 (36%) |
| 5-17 | 296 (72%) | 113 (28%) |
| 18-50 | 123 (70%) | 53 (30%) |
| 51-64 | 45 (75%) | 15 (25%) |
| ≥ 65 | 14 (58%) | 10 (42%) |
| Preexisting conditions |  |  |
| No underlying condition^2^ | 398 (72%) | 153 (28%) |
| Underlying condition^3^ | 182 (69%) | 81 (31%) |
| Missing/Unknown | 5 (63%) | 3 (37%) |

^1^ At higher risk for influenza complications is defined as anyone < 5 years (especially age < 2 years) , **≥** 65 years of age, or who have a preexisting condition defined as increasing the risk for influenza complications [4].

^2-3^Underlying conditions include heart disease, asthma, cancer, diabetes, kidney, obesity, liver, and immune conditions.

**Supplemental Table 3.** Oseltamivir initiation among participants who did or did not seek care within 48 hours of symptom onset by overall risk status, age category, and preexisting conditions in our outpatient cohort of influenza-positive index participants enrolled in the Respiratory Viral Transmission Network.

|  | Sought care within 48 hours of symptom onset | | Did not seek care within 48 hours of symptom onset | |
| --- | --- | --- | --- | --- |
|  | **No oseltamivir (%)** | **Initiated oseltamivir (%)** | **No oseltamivir (%)** | **Initiated oseltamivir (%)** |
| Overall | 322 (55%) | 263 (45%) | 176 (74%) | 61 (26%) |
| At higher risk for influenza complications |  |  |  |  |
| No | 179 (59%) | 125 (41%) | 82 (77%) | 25 (23%) |
| Yes^1^ | 141 (51%) | 137 (49%) | 92 (72%) | 35 (28%) |
| Missing/Unknown |  |  |  |  |
| Age category | **No oseltamivir (%)** | **Initiated oseltamivir (%)** | **No oseltamivir (%)** | **Initiated oseltamivir (%)** |
| < 2 | 21 (50%) | 21 (50%) | 9 (90%) | 1 (10%) |
| 2-4 | 51 (78%) | 14 (22%) | 31 (86%) | 5 (14%) |
| 5-17 | 185 (63%) | 111 (37%) | 87 (77%) | 26 (23%) |
| 18-50 | 51 (41%) | 72 (59%) | 38 (72%) | 15 (28%) |
| 51-64 | 12 (27%) | 33 (73%) | 5 (33%) | 10 (67%) |
| ≥ 65 | 2 (14%) | 12 (86%) | 6 (60%) | 4 (40%) |
| Preexisting conditions |  |  |  |  |
| No underlying condition^2^ | 243 (61%) | 156 (39%) | 120 (79%) | 32 (21%) |
| Underlying condition^3^ | 77 (42%) | 105 (58%) | 53 (65%) | 28 (35%) |
| Missing/Unknown | 2 (67%) | 1 (33%) | 3 (60%) | 2 (40%) |

^1^ At higher risk for influenza complications is defined as anyone < 5 years (especially age < 2 years), **≥** 65 years, or who have a preexisting condition defined as increasing the risk for influenza complications [4].

^2-3^Underlying conditions include heart disease, asthma, cancer, diabetes, kidney, obesity, liver, and immune conditions.
